# Clinical diagnosis of 8274 samples with 2019-novel coronavirus in Wuhan

**DOI:** 10.1101/2020.02.12.20022327

**Authors:** Ming Wang, Qing Wu, Wanzhou Xu, Bin Qiao, Jingwei Wang, Hongyun Zheng, Shupeng Jiang, Junchi Mei, Zegang Wu, Yayun Deng, Fangyuan Zhou, Wei Wu, Yan Zhang, Zhihua Lv, Jingtao Huang, Xiaoqian Guo, Lina Feng, Zunen Xia, Di Li, Zhiliang Xu, Tiangang Liu, Pingan Zhang, Yongqing Tong, Yan Li

## Abstract

**Background:** 2019-Novel coronavirus (2019-nCoV) outbreaks create challenges for hospital laboratories because thousands of samples must be evaluated each day. Sample types, interpretation methods, and corresponding laboratory standards must be established. The possibility of other infections should be assessed to provide a basis for clinical classification, isolation, and treatment. Accordingly, in the present study, we evaluated the testing methods for 2019-nCoV and co-infections.

**Methods:** We used a fluorescence-based quantitative PCR kit urgently distributed by the Chinese CDC to detect 8274 close contacts in the Wuhan region against two loci on the 2019-nCoV genome. We also analyzed 613 patients with fever who underwent multiple tests for 13 respiratory pathogens; 316 subjects were also tested for 2019-nCoV.

**Findings:** Among the 8274 subjects, 2745 (33.2%) had 2019-nCoV infection; 5277 (63.8%) subjects showed negative results in the 2019-nCoV nucleic acid test (non-2019-nCoV); and 252 cases (3.0%) because only one target was positive, the diagnosis was not definitive. Eleven patients who originally had only one positive target were re-examined a few days later; 9 patients (81.8%) were finally defined as 2019-nCoV-positive, and 2 (18.2%) were finally defined as negative. The positive rates of nCoV-NP and nCovORF1ab were 34.7% and 34.7%, respectively. nCoV-NP-positive only and nCovORF1ab-positive cases accounted for 1.5% and 1.5%, respectively. In the 316 patients with multiple respiratory pathogens, 104 were positive for 2019-nCov and 6/104 had co-infection with coronavirus (3/104), influenza A virus (2/104), rhinovirus (2/104), and influenza A H3N2 (1/104); the remaining 212 patients had influenza A virus (11/202), influenza A H3N2 (11/202), rhinovirus (10/202), respiratory syncytial virus (7/202), influenza B virus (6/202), metapneumovirus (4/202), and coronavirus (2/202).

**Interpretation:** Clinical testing methods for 2019-nCoV require improvement. Importantly, 5.8% of 2019-nCoV infected and 18.4% of non-2019-nCoV-infected patients had other pathogen infections. It is important to treat combined infections and perform rapid screening to avoid cross-contamination of patients. A test that quickly and simultaneously screens as many pathogens as possible is needed.

**Funding:** No founding was received

**Research in context:** *Evidence before this study:* We searched PubMed for articles published up to January 31, 2020 using the keywords “2019 novel coronavirus” or “2019-nCoV”. No published study on the characteristics of 2019-nCoV tests or 2019-nCoV co-infections was found. We only noted recent laboratory findings for other tests of patients infected with 2019-nCoV.

*Added value of this study:* Positive detection of nCoV-NP or nCovORF1ab is presented, and individuals with/without 2019-nCoV infections or with inconclusive results were identified. Patients with inconclusive results may be diagnosed with 2019-nCoV infection or found to be negative for the infection after resampling and retesting in the next few days. Approximately 5.8% of the subjects diagnosed with 2019-nCoV had co-infection.

*Implications of all the available evidence:* Management of the population showing inconclusive results should be given attention; additionally, such results can be minimized by improving the sampling, sample pretreatment, and testing methodologies. When diagnosing 2019-nCoV subjects, the possibility of co-infection should be considered. Finally, better clinical detection methods are needed to simultaneously screen as many pathogens as possible.

## INTRODUCTION

An outbreak of novel coronavirus occurred in Wuhan, China in December 2019; since submitting this manuscript, more than 10,000 cases have been confirmed in all provinces of China and 200 deaths have occurred in Wuhan ^1^. Infections spread by air travel have also been reported in Japan, Thailand, Singapore, Republic of Korea, and many other countries^2^. On January 7, 2020, Chinese scientists isolated a novel coronavirus (nCoV) from patients in Wuhan, the genome of which was uploaded to NCBI on January 10^3,4^. Human airway epithelial cells were used to isolate the novel coronavirus, named as 2019-nCoV, which formed another clade within the subgenus sarbecovirus, Orthocoronavirinae subfamily. 2019-nCoV differs from Middle East respiratory syndrome-CoV and severe acute respiratory syndrome-CoV and is the seventh member of the family of coronaviruses that infect humans ^4^. The clinical features of 41 patients infected with 2019 novel coronavirus were described by front-line doctors. It was found that the 2019-nCoV infection caused clusters of severe respiratory illness similar to SARS coronavirus and was associated with intensive care unit admission and high mortality, but had numerous different features from SARS. Among the 41 patients, 6 patients have died since the paper was published^5^. Although the death rate of 2019-nCoV is currently lower than that of SARS, transmission between patients is more prevalent. Clinical evidence showed that one patient infected 14 doctors and nurses. Additionally, in five patients in a family cluster who presented with unexplained pneumonia after returning to Shenzhen, Guangdong province, China, after a visit to Wuhan, an additional family member who did not travel to Wuhan was infected ^6^. This virus can remain in patients for many days, including in some carriers who did not show classic syndromes. This may lead to the infection of many doctors and nurses, as well as rapid dissemination of the virus from Wuhan to many other locations.

The rapid and accurate diagnosis of 2019-nCoV infection in patients and carriers is key for preventing and controlling this epidemic. Thus, fast and accurate clinical detection methods for 2019-nCoV are particularly important. The China CDC recommends the use of a specific real-time RT-PCR method with specific primers and probes to detect the 2019-nCoV open reading frame (ORF1ab) and nucleoprotein (N) gene regions If both targets are positive, the patient is defined as having a laboratory-confirmed infection ^7^.

However, during the testing process, various factors can affect the outcomes of analysis and resulting in false-positive or false-negative results. This makes it difficult to perform clinical diagnosis and control the epidemic situation. In addition, 2019-nCoV may be co-infected with other pathogens. Therefore, patients suspected of having 2019-nCoV infection should also be evaluated by other detection methods to screen for additional respiratory pathogens. However, up-to-date data on co-infected pathogens and their proportions in combination with the 2019-nCoV infection are needed. Our clinical laboratory is located at the center of the epidemic and currently has tested the largest number of samples for 2019-nCoV infection worldwide; we have information for 8274 recent cases (January 21 to February 9, 2020) available for analyzing 2019-nCoV. Determining the laboratory characteristics of 2019-nCoV testing and 2019-nCoV infection with other pathogens will provide a reference for epidemic prevention and clinical treatment in Wuhan and other areas combating this epidemic. Thus, in this study, we evaluated the testing methods for 2019-nCoV, as well as co-infections with other pathogens.

## METHODS

### Sources of Data

Data collected on January 20 to February 9, 2020 at Renmin Hospital of Wuhan University were evaluated. A total of 8397 close contact subjects were enrolled. Of the 8397, 123 patients were subjected to re-sampling (Ct value for the nCoV-NP gene and/or nCovORF1ab gene between 37–40) and were excluded from further analysis. Finally, 8274 subjects (median age 47 years, range 32–62 years) were included in the analysis; men accounted for 37.0% of patients. Nasopharyngeal swab samples were collected from each subject by clinicians, and 63 of 8274 patients (median age 62 years, range 45–69 years, 65.1% male). Sputum samples were collected on the same day. A total of 613 subjects were evaluated by multiple tests for 13 suspected respiratory pathogens during this period (median age 51 years, range 31–63 years, 49.5% male). Of these, 316 (median age 56 years, range 36–66 years, 51.6% male) were also tested for 2019-nCoV simultaneously. The study design was approved by the Ethics Committee of the People’s Hospital of Wuhan University. Data were collected from routine clinical practice, and informed consent was not required.

### Viral Diagnostic Methods

#### Detection of 2019-nCoV

We used the Viral Nucleic Acid Kit (Health, Ningbo, China) to extract nucleic acids from clinical samples according to the kit instructions. A 2019-nCoV detection kit (Bioperfectus, Taizhou, China) was used to detect the ORF1ab gene (nCovORF1ab) and the N gene (nCoV-NP) according to the manufacturer’s instructions using real-time RT-PCR. If the circulation threshold (Ct value) was lower than 37, the gene detection results were considered as positive. If the Ct value was greater than or equal to 40, it was considered that the gene was not present. If the Ct value was between 37 and 40, the presence of the gene was suspected. The clinician was notified to perform resampling and review the results. If the nCovORF1ab and nCoV-NP both showed positive results, the infection was considered as laboratory-confirmed.

#### Detection of 13 types of respiratory pathogens

According to the operating instructions (Health, Ningbo, China), respiratory electrophoresis fragment analysis with PCR was used to screen for infection with 13 respiratory pathogens, including adenovirus, boca virus, influenza A virus, H1N1, H3N2, influenza B viruses, coronavirus, metapneumovirus, parainfluenza virus, respiratory syncytial virus, rhinovirus, mycoplasma pneumoniae, and chlamydia.

### Statistical Analysis

Continuous variables are expressed as the median (interquartile range); categorical variables are expressed as percentages. Kruskal-Wallis H-test was used to compare means between multiple groups, and Dunnett test was used for pairwise comparison. The characteristics of subjects included age and gender. The positive rates of the nCoV-NP and/or nCovORF1ab genes were counted in 8274 subjects. The quantiles were used as a reference. We grouped the patients by gender, and counted the subjects diagnosed with the infection (2019-nCoV group), negative subjects (non-2019-nCoV group), and those with an unclear diagnosis (inconclusive group). We analyzed the infection status of 13 respiratory pathogens in the test subjects and grouped the 2019-nCoV infection status to analyze the infection in the 2019-nCoV group and non-2019-nCoV group. A two-sided α of less than 0.05 was considered as statistically significant. Statistical analyses were conducted using R software, version 3.5.2.

## RESULTS

Of the 8274 subjects, 2745 (33.2%) were diagnosed with the 2019-nCoV infection and classified into the 2019-nCoV group, whereas 5277 (63.8%) were negative for infection according to the 2019-nCoV nucleic acid test and were classified into the non-2019-nCoV group. A total of 252 patients (3.0%), we were temporarily unable to obtain a clear diagnosis because only one target gene was positive. This group of subjects was classified as the inconclusive group. The median ages of subjects in the 2019-nCoV, non-2019-nCoV, and inconclusive groups were 56 years (range, 42–67 years), 40 years (range, 30–57 years), and 52 years (range, 35–64 years), respectively. Dunnett test showed that the age of subjects in the non-2019-nCoV group was significantly lower than in the 2019-nCoV group (t = -20.724, p < 0·001) and inconclusive group (t = -4.251, p < 0·001), whereas the 2019-nCoV group did not significantly differ in age from the inconclusive group (t = -2.218, p = 0.052).

In the non-2019-nCoV group, one case was positive only for nCoV-NP, when they were first evaluated, whereas both nCovORF1ab and nCoV-NP were negative after resampling and revaluation on the fourth day. One case were positive only for nCovORF1ab when they were first evaluated, whereas both nCovORF1ab and nCoV-NP were negative after resampling and revaluation on the third day.

In the 2019-nCoV group, two subjects was positive only for nCovORF1ab; same-day re-sampling showed the same results, whereas re-sampling on the next day and third day respectively for nCovORF1ab and nCoV-NP revealed positive results. Six subjects was positive only for nCovORF1ab, which was confirmed by re-sampling on the fourth day (range, 1-10). One case was positive only for nCoV-NP; these results were confirmed by re-sampling and re-examination on the third day. Forty-two subjects was negative both for nCoV-NP and nCovORF1ab, these results were confirmed by re-sampling and re-examination 5.5 days (range, 2.5-7.0) later.

The positive rates of nCoV-NP and nCovORF1ab in the tested population were 34.7% and 34.7%, respectively. Only 1.5% cases were positive for nCoV-NP and 1.5% were positive for nCoVORF1ab. 33.2% met the 2019-nCov diagnostic criteria (nCoV-NP and nCovORF1a were positive simultaneously). As shown in Table 2.

**Table 1.** Characteristics of individuals with/without 2019-nCoV Infected in Wuhan.

| Characteristic | Non-2019-nCoV<br>(n=5277) | 2019-nCoV<br>(n=2745) | Inconclusive<br>(n=252) |
| --- | --- | --- | --- |
| Male sex-no.total no. (%) | 1542/3845 (40.1) | 882/1881 (46.9) | 61/135 (45.2) |
| Median age (rang)-yr | 40 (30-57) | 56 (42-67) | 52 (35-64) |
| <15 yr | 60/3847 (1.6) | 4/1867 (0.2) | 1/128 (0.8) |
| 15-44 yr | 2068/3847 (53.8) | 528/1867 (28.3) | 46/128 (35.9) |
| 45-64 yr | 1127/3847 (29.3) | 773/1867 (41.4) | 50/128 (39.1) |
| ≥65 yr | 592/3847 (15.4) | 562/1867 (30.1) | 31/128 (24.2) |

Inconclusive: either nCoV-NP or nCovORF1ab was positive (not all were positive)
Reduced denominators indicate missing data. Percentages may not total 100 because of rounding.

**Table 2.** Positive results in 2019-nCov test and confirmed diagnosis rate.

|  | nCov-NP (n=8274) | nCovORF1ab (n=8274) |
| --- | --- | --- |
| Positive-no.total no. (%) | 2873/8274 (34.7) | 2867/8274 (34.7) |
| nCov-NP (+) and nCovORF1ab (-) | 128/8274 (1.5) | - |
| nCov-NP (-) and nCovORF1ab (+) | - | 124/8274 (1.5) |
| nCov-NP (+) and nCovORF1ab (+) | 2745/8274 (33.2) |  |

The tested population was divided into two groups by gender. The number of patients diagnosed with 2019-nCoV in each age group is shown in Figure 1A. Of the 2485 male subjects, 882 (35.5%) were diagnosed as having 2019-nCov infection and 61 cases (2.5%) could not be clearly diagnosed. Of the 3376 female subjects, 999 (29.6%) were diagnosed with 2019-nCov infection and 74 (2.2%) could not be determined (Figure 1B).

**Figure 1.**
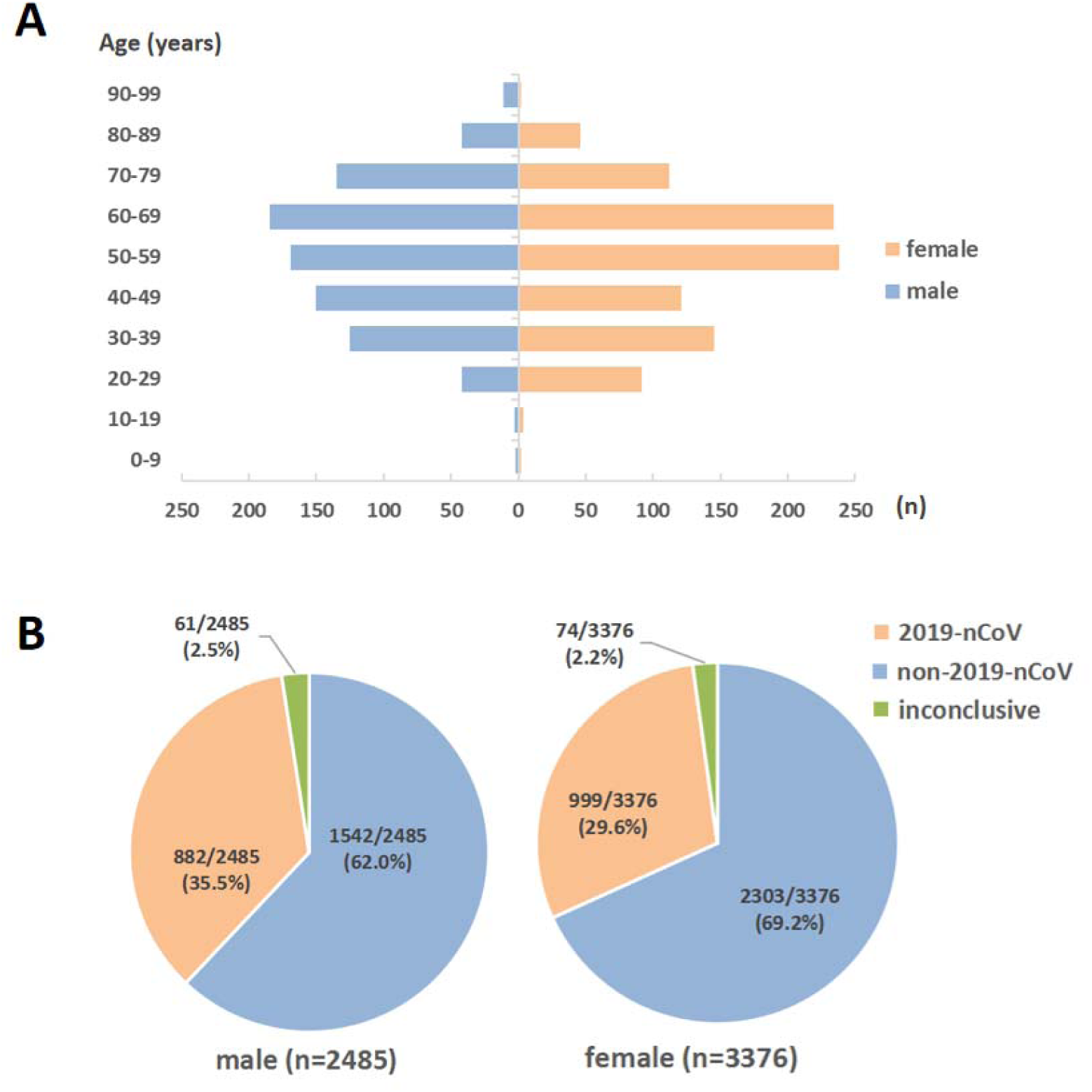
Relationship between 2019-nCov diagnosis rate and age and gender of patients. (a: The number of patients diagnosed with 2019-nCoV in each age group; b: Test population 2019-nCoV (relationship between 2019-nCoV diagnosis rate and gender).

Sixty-three cases of nasopharyngeal swab samples showed clear diagnoses; 2 cases of sputum samples from the corresponding subjects could not be judged, and 1 case of sputum samples diagnosed as negative was evaluated by collecting nasopharyngeal swabs and confirmed as positive for 2019-nCoV(Table 2).

**Table 2.** Comparison of diagnosis rate of nasopharyngeal swab samples and sputum samples.

|  |  | Nasopharyngeal swab (n=63) |  |  | Total |
| --- | --- | --- | --- | --- | --- |
|  |  | laboratory-confirmed | negative | suspicious |  |
| Sputum<br>(n=63) | laboratory-confirmed | 6 | 0 | 0 | 6 |
|  | negative | 1 | 54 | 0 | 55 |
|  | suspicious | 1 | 1 | 0 | 2 |
|  | Total | 8 | 55 | 0 | 63 |

Suspicious: Ct value was 37–40. Resampling is recommended.

Of the 613 subjects tested for the nucleic acids of 13 respiratory pathogens, 14.2% (45/316) of subjects had a pathogen infection. The positive rates of each pathogen were as follows: influenza A virus 5.55% (34/613), rhinovirus 4.73% (29/613), influenza A H3N2 4.57% (28/613), respiratory syncytial virus 4.40% (27/613), influenza B virus 4.08% (25/613), coronavirus 2.12% (13/613), metapneumovirus 1.63% (10/613), H1N1 0.65% (4/613), adenovirus 0.65% (4/613), mycoplasma pneumoniae 0.49% (3/613), parainfluenza virus 0.49% (3/613), chlamydia 0.49% (3/613), and boca virus 0.33% (2/613) (Figure 2a).

**Figure 2.**
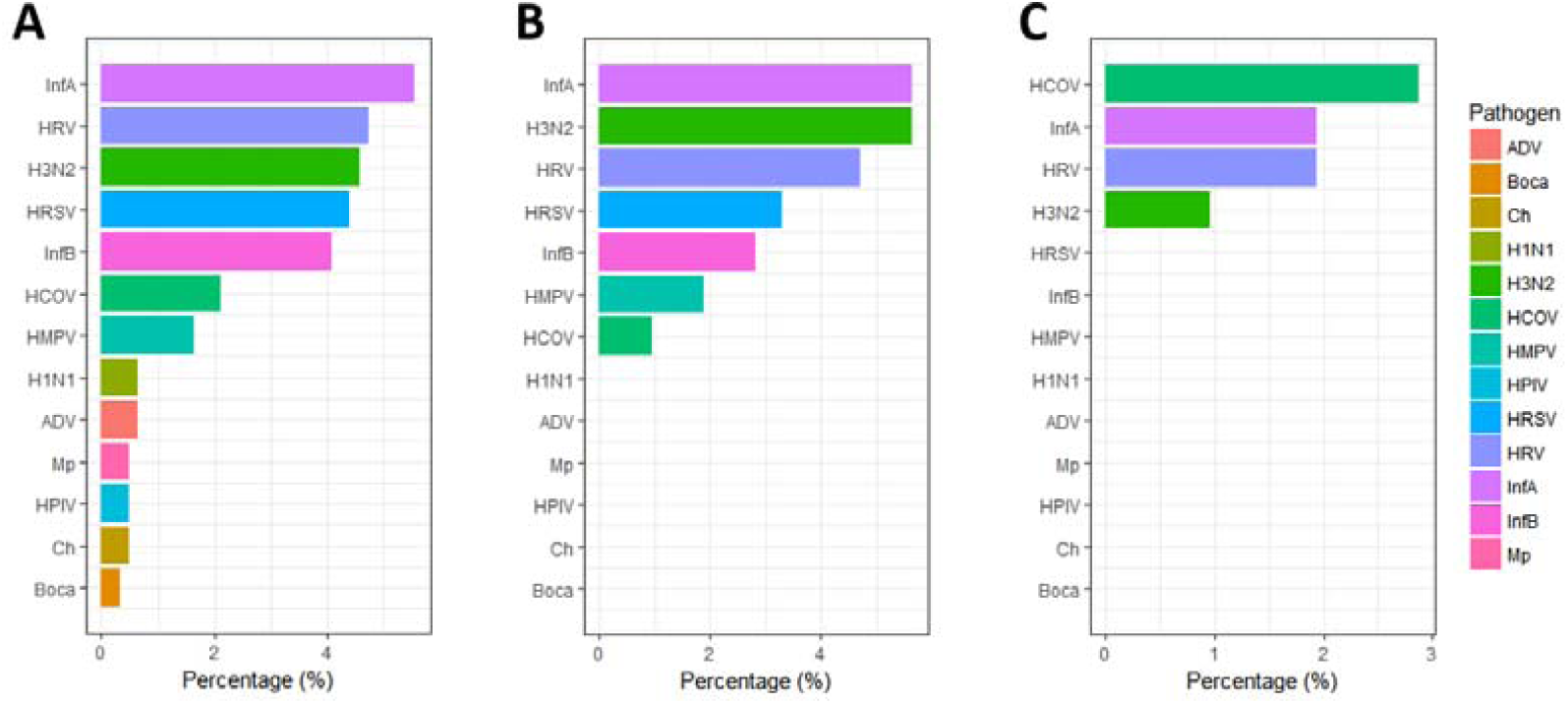
Comparison of 2019-nCov infection rates with infection rates of 13 other respiratory pathogens. (a: Pathogen-positive rates for all 13 respiratory pathogens tested, n = 614; b: 2019-nCov-negative subjects, 13 respiratory-pathogen-positive rates, n = 202; c: 2019-nCov-confirmed subjects. Positive rate of 13 respiratory pathogens in the test population, n= 104. ADV: Adenovirus; Boca: Boka virus; Ch: Chlamydia; H1N1: H1N1 Influenza A virus; H3N2: H3N2 Influenza A virus; HCOV: Human coronavirus; HMPV: Human metapneumovirus; HPIV: Human parainfluenza virus; HRSV: Respiratory syncytial virus; HRV: Human rhinovirus; InfA: Influenza A virus; InfB: Influenza B virus; Mp: *Mycoplasma pneumoniae*).

Among the subjects who were 212 2019-nCov-negative, 18.4% (39/212) were infected with pathogens. The nucleic acid-positive rates of the 13 respiratory pathogens were as follows: influenza A virus 5.66% (11/202), A H3N2 5.66% (11/202), rhinovirus 4.72% (10/202), respiratory syncytial virus 3.30% (7/202), influenza B virus 2.83% (6/202), metapneumovirus 1.89% (4/202), and coronavirus 0.94% (2/202) (Figure 2B).

Among the 104 patients diagnosed with 2019-nCov, 5.8% (6/104) had co-infection. The nucleic acid positive rates of the 13 respiratory pathogens were as follows: coronavirus 2.88% (3/104), influenza A virus 1.94% (2/104), rhinovirus 1.94% (2/104), and influenza A H3N2 0.96% (1/104) (Figure 2C).

## DISCUSSION

Here, we used 8274 samples to initially evaluate the laboratory testing characteristics of 2019-nCoV. In clinical testing, although most subjects showed a clear diagnosis, other subjects did not. This is not conducive for treating potential infections and controlling the 2019-nCoV epidemic. In addition, 2019-nCoV may be co-infected with other pathogens; however, the types of co-infected pathogens and the proportion of 2019-nCoV infection is unclear; whether these co-infection conditions require improvements in current treatments is unknown.

The results of this study indicate that of the 8274 subjects evaluated by the 2019-nCoV nucleic acid test, 2745 (33.2%) were confirmed to be infected with 2019-nCoV, whereas 5277 (63.8%) showed negative results. For 252 cases (3.0%), we were temporarily unable to obtain a clear diagnosis because only one target gene was positive. The proportion of males infected (35.5%) was higher than that of females (29.6%). Among the 104 patients who tested positive for 2019-nCov, 5.8% were co-infected with other pathogens. Our findings provide important data for clinical testing, clinical diagnosis, and treatment, as well as for 2019-nCoV epidemic control.

### Clinical test

The results of test in which only one target gene was positive requires further attention. From the perspective of clinical testing, this may have occurred because of sample collection problems. Poor sample quality greatly affects clinical testing results. We compared sputum samples with nasopharyngeal swab samples from 63 subjects collected on the same day. The results obtained from the 63 nasopharyngeal swab samples were very clear, whereas sputum samples showed a Ct value of 37–40 and some-false negatives. This may be because sputum is not a homogeneous substance. When extracting nucleic acids, the sampling site of the sputum sample can affect the quality and quantity of 2019-nCoV in the extracted nucleic acid, in turn affecting the test results. In addition, the properties and position of the sputum in the trachea vary widely. This can prevent effective control of the sample quality of the sample during the sampling process, which can affect the test results, particularly in patients at an early stage of infection when the virus has not yet multiplied. Second, we used pharyngeal swabs which are flexible and change shape as the nasopharynx bends; thus, samples can be obtained from deeper locations, which is helpful for collecting samples with a higher virus content. Additionally, the 2019-nCoV epidemic is currently a serious public health threat, and thus personnel with rich experience in clinical sampling should be designated to collect samples. Finally, attention should be paid to environmental pollution during sampling, and hospital management departments should set up special sampling rooms and management measures to prevent the samples from being contaminated with 2019-nCoV in the environment.

Problems related to sample pre-processing should also be considered. Before clinical testing of the samples, steps such as nucleic acid extraction are required. Experimental operators and management personnel should pay attention to whether the laboratory is qualified for testing. The laboratory environment and its equipment should have the ability to prevent cross-contamination of samples, prevent occupational exposure of personnel, and be equipped with comprehensive disinfection equipment and emergency measures for occupational exposure. Only nCovORF1ab showed positive results during evaluation, and the two targets were negative after re-sampling and re-examination on the next a few days; thus, contamination may have occurred. Contamination may arise from the sampling or processing process. Additionally, personnel familiar with the experimental skills required should perform the analysis. Finally, the 2019-nCoV test kit should meet the clinical requirements.

Regarding detection problems, inspectors and managers should determine whether the kit used meets the required standards, reagents are stored properly, and equipment used has aging parts.

For the pathological process of virus infection, we observed a case in which only nCovORF1ab-positive subjects were resampled on the same day and tested, revealing consistent results. Positive results were observed for CoV-NP and nCovORF1ab, which was confirmed by resampling on the next day. Therefore, an unclear diagnosis may be related to the pathological stage of the patient.

Bacterial proteins such as those produced by *Streptococcus pneumoniae* and *Staphylococcus aureus* can promote the spread of influenza virus^8^. Therefore, from a mechanistic perspective, patients with non-2019-nCoV-induced pneumonia are more likely to be infected with 2019-nCoV than healthy people. In contrast, people infected with 2019-nCoV (particularly severely ill patients) will also have secondary infections. Nanshan Chen et al. studied 99 patients with 2019-nCoV infection and found that 1 (1%) was infected with bacteria and 4 (4%) were infected with fungi^9^. These issues demonstrate that new requirements are needed for clinical testing. In this study, we tested for the presence of 13 types of respiratory pathogens to detect non-bacterial infections; however, to detect infection with bacterial pathogens, medical institutions generally use traditional smears and cultures. It is well known that the sputum smear-positive rate is low, and the culture process is very time-consuming. Therefore, a broad-spectrum, rapid, and accurate bacterial screening and detection platform (even for most known and unknown pathogens) is needed in the clinic to identify pathogens, judge the patient’s condition, and avoid blind administration of drugs.

### Clinical diagnosis and treatment

The median age of the 2019-nCoV group in this study was 56 years (range 42–67), which was close to the first 425 patients diagnosed (59 years, range 15–89 years) ^10^. Interestingly, even if a clear diagnosis cannot be obtained for the inconclusive group, the median age of these patients (52 years, range 35–64 years) was very similar to that of the 2019-nCoV group (56 years, range 42–67 years). These patients were much older than patients in the non-2019-nCoV group (40 years, 30–57 years). Thus, 2019-nCoV infection is likely to be undetected in subjects in the inconclusive group. We observed that in several subjects for whom a clear diagnosis could not be initially obtained, 2019-nCoV infection was confirmed after re-sampling and re-evaluation on the following days. However, the predictions based on the age variable alone are well-supported. Moreover, two patients for whom a clear diagnosis could not be initially obtained were re-sampled on the next a few days, showing nucleic acid-negative test results. Although this may have occurred because of contamination during sampling and testing, the presence of this group indicates that the inconclusive group may not have undetected 2019-nCoV infection. Further studies are needed to evaluate this point.

Screening for 13 respiratory pathogens revealed that 2019-nCoV can be co-infected with other viruses, mainly coronavirus 2.88% (3/104), influenza A virus 1.94% (2/104), rhinovirus 1.94% (2/104), and A3 H3N2 0.96% (1/104). Therefore, while treating 2019-nCoV, other viruses should also be treated. In addition, 2019-nCoV is secondary to infection with bacteria and fungi^11^. Common secondary infections include *Acinetobacter baumannii, Klebsiella pneumoniae, Aspergillus flavus, Candida glabrata*, and *Candida albicans*. Infection of some strains that are prone to drug resistance (such as *A. baumannii*) make treatment more difficult and patients may develop septic shock^12^. Our results indicate that patients infected with 2019-nCoV are mostly middle-aged and elderly, and their resistance to infection is generally low; thus, prophylactic antibiotics can reduce their mortality and comorbidity.

### Outbreak control

Our results show that 3.0% of subjects were positive for only one target gene in initial testing; re-sampling and review after several days revealed both diagnosed and nucleic acid-negative cases. Because of the limited number of samples, we could not determine whether false-positive or false-negative results were more likely; however, those who are positive for only one target gene in initial testing should be isolated and observed as much as possible to avoid potential 2019-nCoV infected persons from spreading the epidemic. Patients infected with 2019-nCoV may be infected with other pathogens which are community-based, iatrogenic, or both, for which precautions should be taken to reduce the risk of cross-infection among patients in the hospital. For example, patients infected with 2019-nCoV should be isolated from those with pneumonia caused by other pathogens. This isolation should involve treatment and monitoring from the patient’s entry to the outpatient clinic to final discharge.

In summary, we analyzed data from 8274 laboratory tests and determined the laboratory test characteristics of 2019-nCoV. We found that 3.0% of subjects could not be clearly diagnosed with 2019-nCoV infection. they may be 2019-nCoV People with nCoV infection may also be uninfected people. Thus, the management of this population should be considered. The generation of such results can be minimized by improving sampling, sample preparation, and testing methods. In addition, 5.8% of the subjects diagnosed with 2019-nCoV were infected with other pathogens. The possibility of co-infection should also be considered; additionally, better clinical detection methods are necessary to simultaneously screen for as many pathogens as possible.

## Data Availability

The data that support the findings of this study are available from the corresponding author on reasonable request.

## Author Contributions

MW and YL had roles in the study design, patient recruitment, data collection, data analysis, data interpretation, literature search, and writing of the manuscript. QW, W-ZX, BQ, J-WW, H-YZ, S-PJ, J-CM, Z-GW, Y-YD, F-YZ, WW, YZ, Z-HL, JJH, X-QG, LF, ZE-X, DL, had roles in the experiments, data collection, data analysis, and data interpretation. Z-LX, T-GL, P-AZ, Y-QT and YL contributed to critical revision of the manuscript. All authors contributed to data acquisition, data analysis, data interpretation, and reviewed and approved the final version.

## Acknowledgments

No founding was received. We thank all clinicians and inspectors at the front line of the 2019-nCoV epidemic.

